# Retrospective Analysis of Clinical Features in 101 Death Cases with COVID-19

**DOI:** 10.1101/2020.03.09.20033068

**Authors:** Hua Fan, Lin Zhang, Bin Huang, Muxin Zhu, Yong Zhou, Huan Zhang, Xiaogen Tao, Shaohui Cheng, Wenhu Yu, Liping Zhu, Jian Chen

**Author notes:** Hua Fan; Lin Zhang; Bin Huang; Muxin Zhu; Yong Zhou have contributed equally to this work. Contributed equally (Hua Fan, Lin Zhang, Bin Huang, Muxin Zhu, Yong Zhou). Joint corresponding authors, **Corresponding Author:** Jian Chen, No.1 Tiane Road, Shu Shan District, The First Affiliated Hospital of USTC, Hefei, Anhui 230036, China, Wenhu Yu, No. 29 Mianzhou road, Xiantao, Hubei 433000, China, Liping Zhu, Wuhan Jinyintan Hospital, No.1 Yintan Road,Dongxihu District, Wuhan, Hu bei430023, China.

## Abstract

**Background:** In December 2019, viral pneumonia outbreaks caused by the novel coronavirus occurred in Wuhan, China, and spread rapidly worldwide, the illness progress of coronavirus disease 2019(COVID-19) in partial patients is rapid and the mortality rate is relatively high. The present study aims to describe the clinical features in the death cases with COVID-19.

**Methods:** In this single center and observational study, we recruited 101 death cases with COVID-19 from Dec 30, 2019 to Feb 16, 2020 in Intensive Care Unit (ICU) of Wuhan Jinyintan Hospital. Demographics, underlying diseases, X-ray/CT images, possible therapy strategies and test results were collected and analyzed on patients admission to the ICU and 48 hours before deaths.

**Findings:** Of 101 COVID-19 dead cases, 47 patients went directly to the ICU because of critical condition, and 54 patients were transferred to ICU with aggravated condition. Over fifty six percent (56.4%) of patients were laboratory confirmed by RT-PCR, and 43.6% were consistent with clinical diagnostic criteria.Among them, 64 were males and 37 were females, with average age of 65.46 years (SD 9.74). Interestingly, all deaths shwed significant difference in blood type distribution, with 44.44% of type A, 29.29% of type B, 8.08% of type AB, and 18.19% of type O.The clinical manifestations of the novel coronavirus pneumonia are non-specific,the common symptoms included fever (91,,90.10%), cough (69,68.32%) and dyspnea (75,74.26%). Neutrophils, procalcitonin(PCT),C-reactive protein(CRP),and interleukin-6(IL-6), D-dimer gradually increased with progress of the disease. Myocardial enzymes were abnormal in most patients at admission, myocardial damage indicators were significantly increased. Sixty one (60.40%) patients were given antiviral drugs, 59(58.42%),received glucocorticoids, 63.37% were given intravenous immunoglobulins, and 44.55% were treated with thymosin preparations. All patients received antibiotic treatment, 63(62.38%) were given restricted antibiotics, 23(22.78%) were administrated to antifungal drugs. Non-invasive ventilator or high-flow oxygen therapy were given in 84(83.17%) patients, and invasive mechanical ventilation was used in 76.24% patients. The median time from acute respiratory distress syndrome(ARDS) to invasive mechanical ventilation was 3.00 days(IQR 0.00-6.00). The duration of invasive mechanical ventilation was 5 days (IQR2.00-8.00).

**Interpretation:** Critical COVID-19 can cause fatal respiratory distress syndrome and multiple organ failure with high mortality. The blood group distribution of the deaths remarkably differred from that of Han population in Wuhan.Heart may be the earliest damaged organ except the lungs. Hospital-acquired pneumonia(HAP) in the later period is worthy of attention.

**Funding:** Supported by “the Fundamental Research Funds for the Central Universities (WK9110000037)”.

## 1. INTRODUCTION

Viral pneumonia outbreaks caused by novel coronavirus occurred in Wuhan, China in December 2019, and spread rapidly worldwide^[1-5]^. The illness progress of partial patients wasrapid and the mortality was found to be 3.4%. By 5^th^ March, 2020, the cumulative number of infections worldwide reached 95,333 cases, of which 3,282 died^[5]^. In 30th January, 2020, the World Health Organization issued a global warning about the highly contagious virus^[6]^, which was named as coronavirus disease 2019 (COVID-19) in 11^th^ February^[7]^. Current studies^[8-10]^ demonstrated that COVID-19 often occured in elderly men with underlying diseases based on the laboratory and clinical features. In early stage, fever and dry cough with reduced number of lymphocytes were the major manifestations, fatal respiratory distress syndrome and multiple organ failure were advanced in the late stage.

However, there have been few studies on the clinical characteristics of death cases due to the small sample size. To understand the clinical characteristics, a retrospective analysis of clinical features in 101 death cases in Wuhan Jinyintan Hospital was carried out.

## 2. METHODS

### 2.1. Study Population

This study collected death cases with COVID-19 from Dec 30, 2019 to Feb 16, 2020 in ICU of Wuhan Jinyintan Hospital. The Hospital specialize for infectious diseases, prescribed by Chinese government as one of the first designated treatment units for patients with the disease. The diagnosis of confirmed and clinical cases was made following Diagnosis and Treatment of Novel Coronavirus Pneumonia (trial version 5)^[11]^. The confirmed cases were those who had pathogenic evidence, positive RT-PCR or highly homologous gene sequencing with known coronavirus. The clinical cases were those who had epidemic history, or clinical symptoms and imaging characteristics. Epidemic history means that the suspected cases has travelling history in Wuhan and surrounding areas within 14 days, or exposed to other infections, communicated with patients with fever or respiratory symptoms from Wuhan and surrounding areas or from case-reporting communities, or clustering onset. The clinical features include fever or respiratory symptoms, and decreased lymphocyte or leukocyte count in the early stage.

This study has been approved by the Ethics Committee of Wuhan Jinyintan Hospital (KY-2020-28.01), and all relevant personnel exempts from informed consent due to the particularity of the disease outbreak.

### 2.2. Data Collection

The retrospective analysis was based on the case reports, nursing records, test results, and imaging results. Data include demographics, underlying diseases, X-ray/CT results, possible therapy strategies and test results of patients on admission to ICU or 48 h before deaths. The therapy strategy represents antiviral and antibacterial treatments, corticosteroid treatment, immunotherapy, and respiratory therapy. The onset time was defined as the day symptom appeared. Records began from onset to admission, admission to ICU, ICU periods, and onset to deaths.

### 2.3. Statistical Analysis

We compared the differences in epidemiologic, clinical, and laboratory measures among patients died of COVID-19 at onset to admission, admission to ICU and 48h before deaths.We present continuous measurements as mean (SD) if they were normally distributed or median (IQR) if not, and categorical variables were described as frequency rates and percentages(%). For laboratory results, we also assessed whether the measurements were outside the normal range. SPSS (version 24.0) was used for all analyses.

## 3. RESULTS

### 3.1. Demographics

By Feb 16, 2020, 101 cases died of COVID-19 in ICU of Wuhan Jinyintan Hospital, among them 57 (56.44%) patients were confirmed by RT-PCR, and 44 (43.6%) were consistent with clinical diagnostic criteria. In the death cases, 47 went directly to the ICU because of critical condition, and 54 were transferred to ICU with aggravated condition. All cases died in the end. Among them, 1 patient died of acute coronary syndrome, while the rest died of respiratory failure and multiple organ failure caused by COVID-19. The cases included 64 males and 37 females, with the age range of 24-83 and average age of 65.46 years (SD 9.74). The majority had underlying diseases with hypertension(42.57%), diabetes (22.77%), neurological disease (9.90%), malignant tumor (4.95%), and respiratory disease(4.95%). The cases with more than two underlying diseases reached 25.74%. In detail, 10 patients had contact history in South China Seafood Market, and 10 patients had close contact history with infections. The blood type distribution was significantly different, with type A (44.44%), type B (29.29%), type AB (8.08%) and type O (18.19%). The median time from onset to hospital was 11.00 days (IQR8.00-13.50). The median time from onset to death was 21.00 days (IQR17.00-27.5)(Table 1).

**Table1.** Demographics of 101 patients with COVID-19

|  | Patients/days(n=101) |
| --- | --- |
| Age,years |  |
| Mean(SD) | 65.46±9.74 |
| Range | 24-83 |
| ≤44 | 3 ( 3.00% ) |
| 45-59 | 19 ( 18.80% ) |
| 60-74 | 60 ( 59.40% ) |
| ≥75 | 19 ( 18.80% ) |
| Sex |  |
| Female | 37 ( 37.00% ) |
| Male | 64 ( 64.00% ) |
| Days of illness |  |
| Days from onset to admission | 11.00 ( 8.00-13.50 ) |
| Days in general ward | 0.00 ( 0.00-4.00 ) |
| Days in ICU | 8.00 ( 4.00-11.00 ) |
| Days from onset to death | 21.00 ( 17.00-27.50 ) |
| Blood group |  |
| A | 44(44.44%) |
| B | 29(29.29%) |
| AB | 8(8.08%) |
| O | 18(18.19%) |
| Exposure history |  |
| Long-term exposure history Huanan seafood market | 7(6.93%) |
| Short-term exposure history Huanan seafood market | 3(2.97%) |
| Casual contact with a patient with COVID-19 | 10(9.90%) |
| Positive RT-PCR | 57(56.44%) |
| Chronic medical illness |  |
| Cardiovascular and cerebrovascular disease | 49 ( 48.51% ) |
| Endocrine system disease | 23 ( 22.77% ) |
| Malignant tumour | 5 ( 4.95% ) |
| Nervous system disease | 10 ( 9.90% ) |
| Respiratory system disease | 5 ( 4.95% ) |

Values are numbers (percentages) or Median (IQR) unless stated otherwise. Percentages do not total up to 100% owing to missing data.

### 3.2. Clinical Features

The relative frequencies of all reported symptoms at the time of admission are shown in Table 2. The most common symptom was fever (91,90.10% of 101 patients), but most patients presented normal temperature after 1-3 days of admission, which may be related to hormone use. Cough (69 [68.32%]) and dyspnea (75 [74.26%]) were also common. Additionally, 31(30.69%) patients had white sputum in the early stage, and 3 patients showed yellowish purulent sputum. Fifty(49.50%) patients showed acute respiratory distress syndrome(ARDS) on admission, and the median time from onset to ARDS was 12 days(IQR9.00-14.00). Other common symptoms included myalgia, general weakness, dizziness, headache, nausea and vomiting, among them 86(85.15%) had more than two symptoms. Twenty one (20.79%) patients had no respiratory symptoms, and 7(6.93%) showed nausea and vomiting. Distinctive from Severe acute respiratory syndrome (SARS), only 2 patients with COVID-19 had diarrhea.

**Table2.**
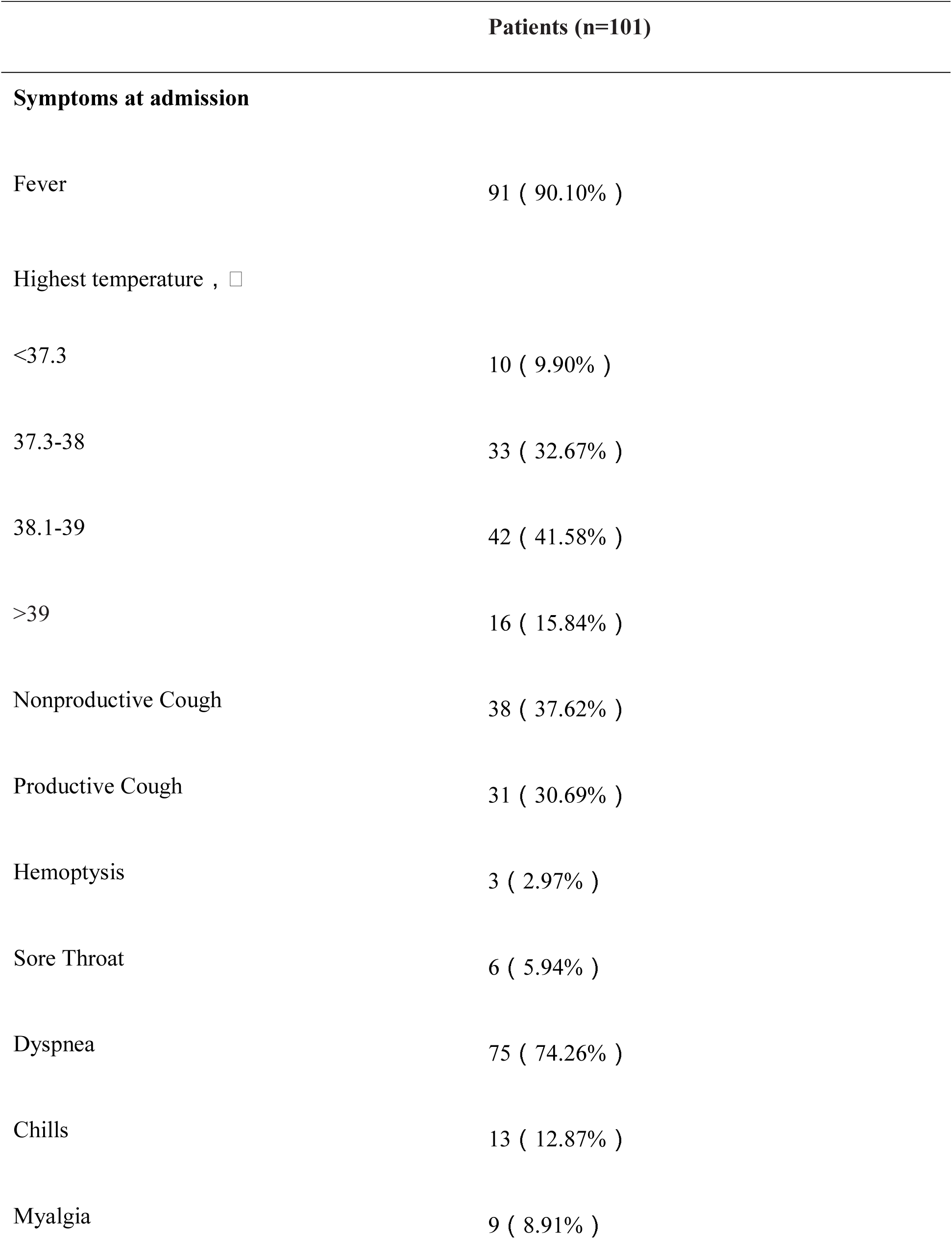
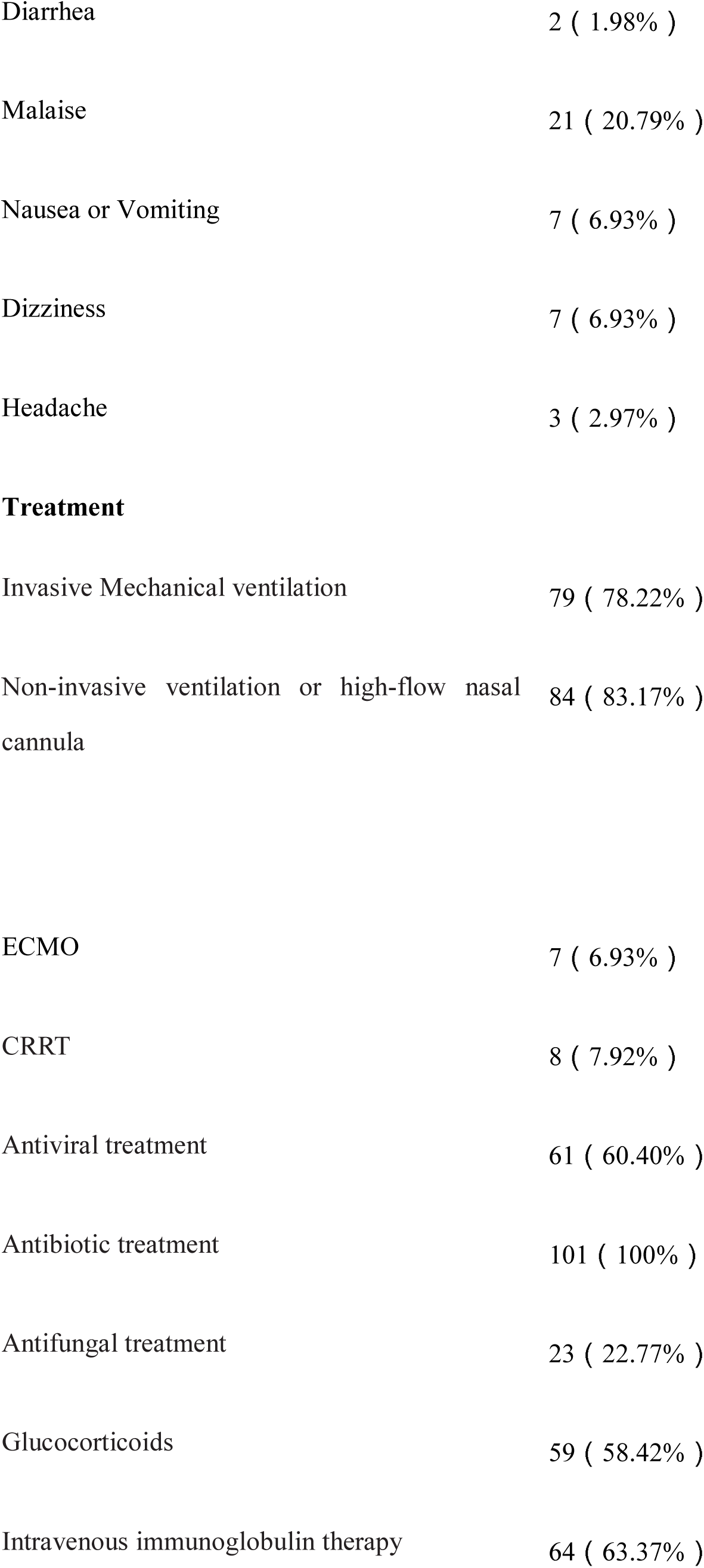
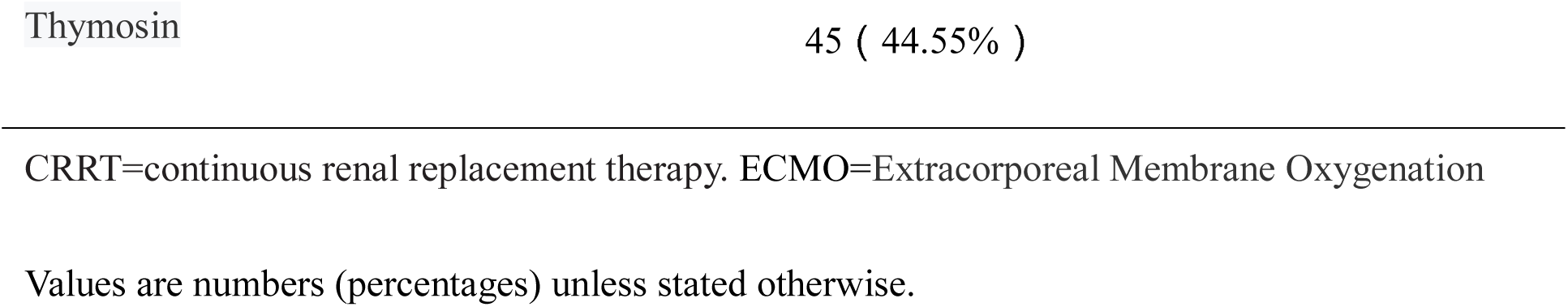
Clinical characteristics and treatment of patients with COVID-19

### 3.3. Radiographic Findings

Most of patients (99 [98.02%] of 101 patients) had abnormal lung imaging at the time of initial admission, 6(5.94%) patients had single-lung and 93(92.08%) had double-lung abnormalities. The disease initially showed scattered plaque-like ground glass in the lungs. Dynamic imaging showed progressive multi-spot patchy shadows in both lungs. Pneumothorax or subcutaneous emphysema occurred in 3 (2.97%) patients.

### 3.4. Laboratory Findings

Blood routine, biochemical reports and inflammation of cases during admission to ICU and 48h before deaths were collected. Most patients showed increased numbers of leukocytes and neutrophils, and decreased lymphocytes count at time of consultation.The inflammatory indicators such as neutrophils, procalcitonin(PCT),C-reactive protein(CRP), and interleukin-6(IL-6) gradually increased as time went on. It was found that the count of patient’s platelets(PLT) decreased, D-dimer and Prothrombin time(PT) increased correspondingly during disease progression by dynamic analysis of coagulation-related indicators. Myocardial enzymes were abnormal in most patients at admission(83 [82.18%] of 101 patients). With the progress of the disease, myocardial damage indicators were significantly increased. It also indicated that there was a high proportion of myocardial damage in the early stage. We also found that liver and kidney damages were not significant at the time of admission and when transferred to ICU, and it significantly turned to be worse 48h before deaths. Experiment data can be seen in Table 3-5 and Figure1.

**Figure1.**
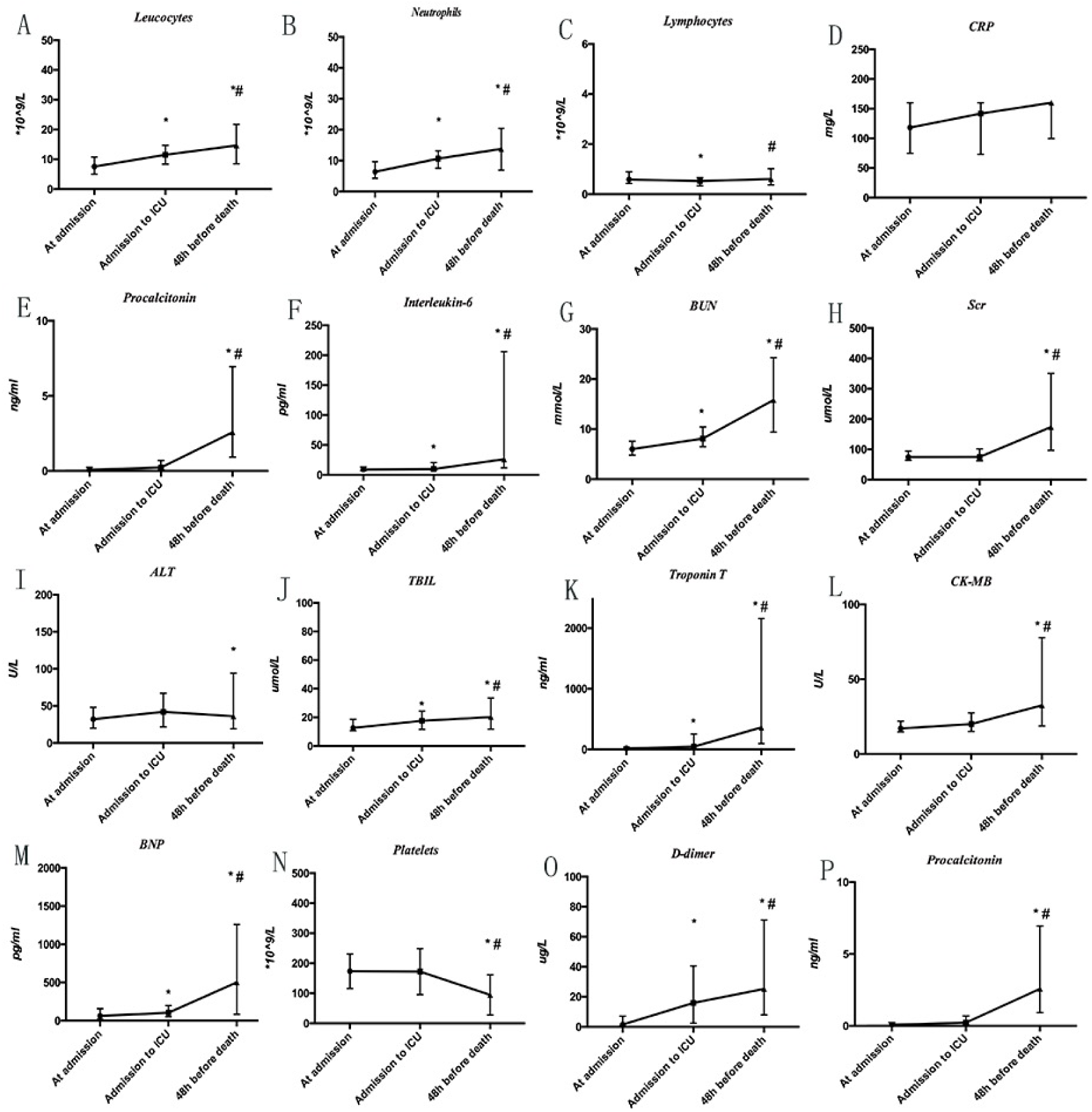
Dynamic Profile of Laboratory Parameters in 101 Patients With COVID-19. Timeline charts illustrate the laboratory parameters in 101 patients with COVID-19 when their entrance into admission, ICU and 48 h before death. \**P* < .05 vs. At admission group, #*P* < .05 vs. Admission to ICU group.

**Table3.**
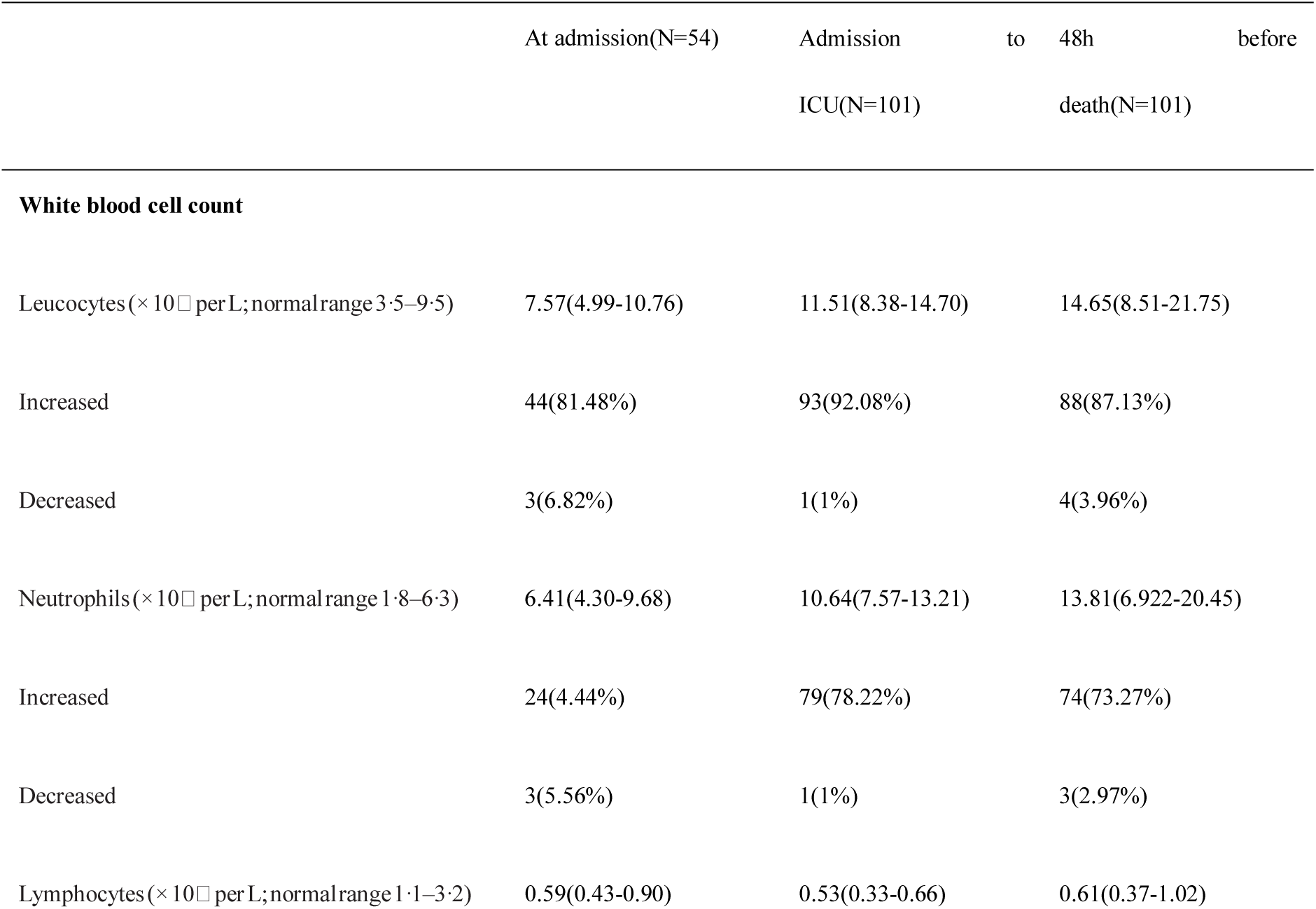
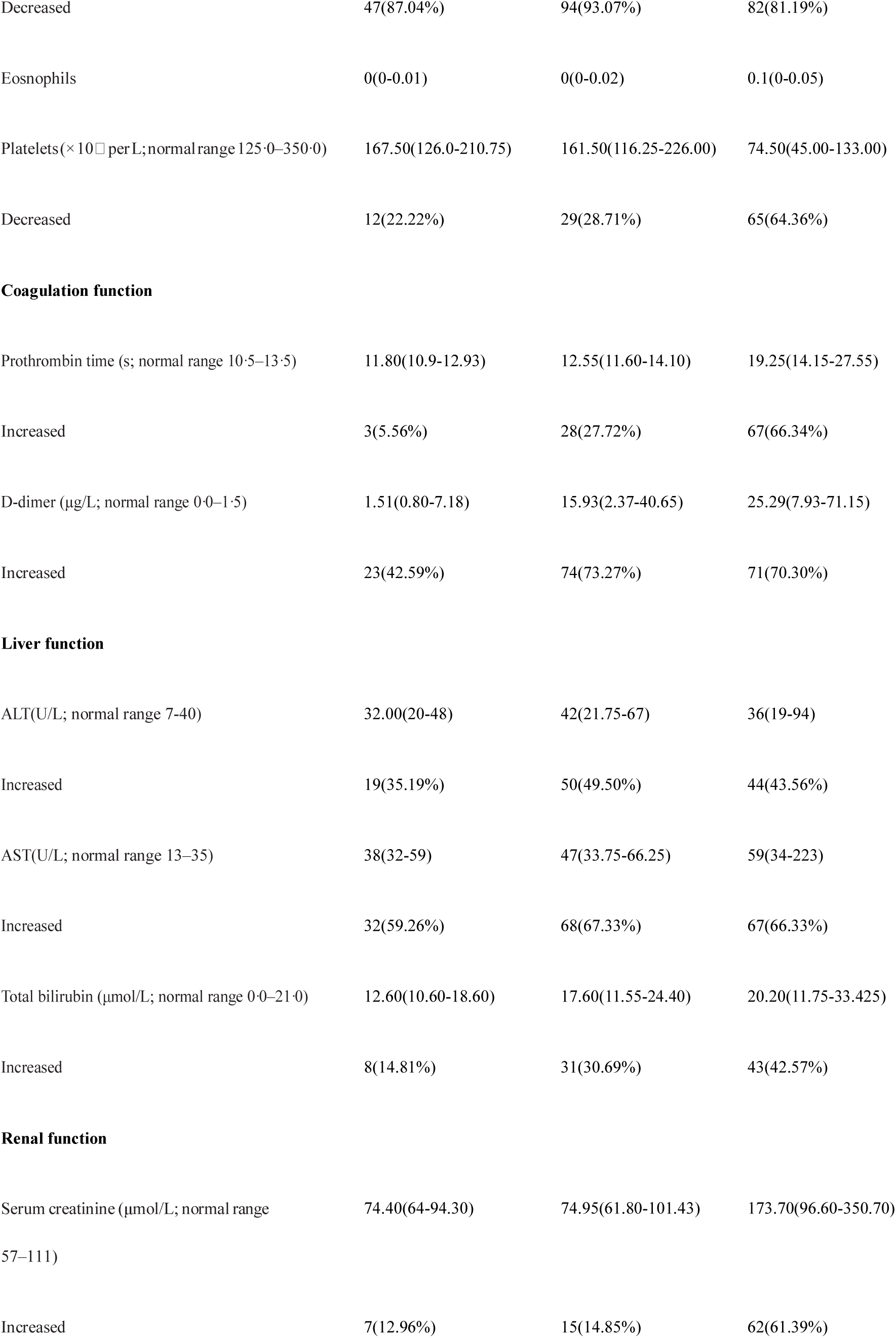
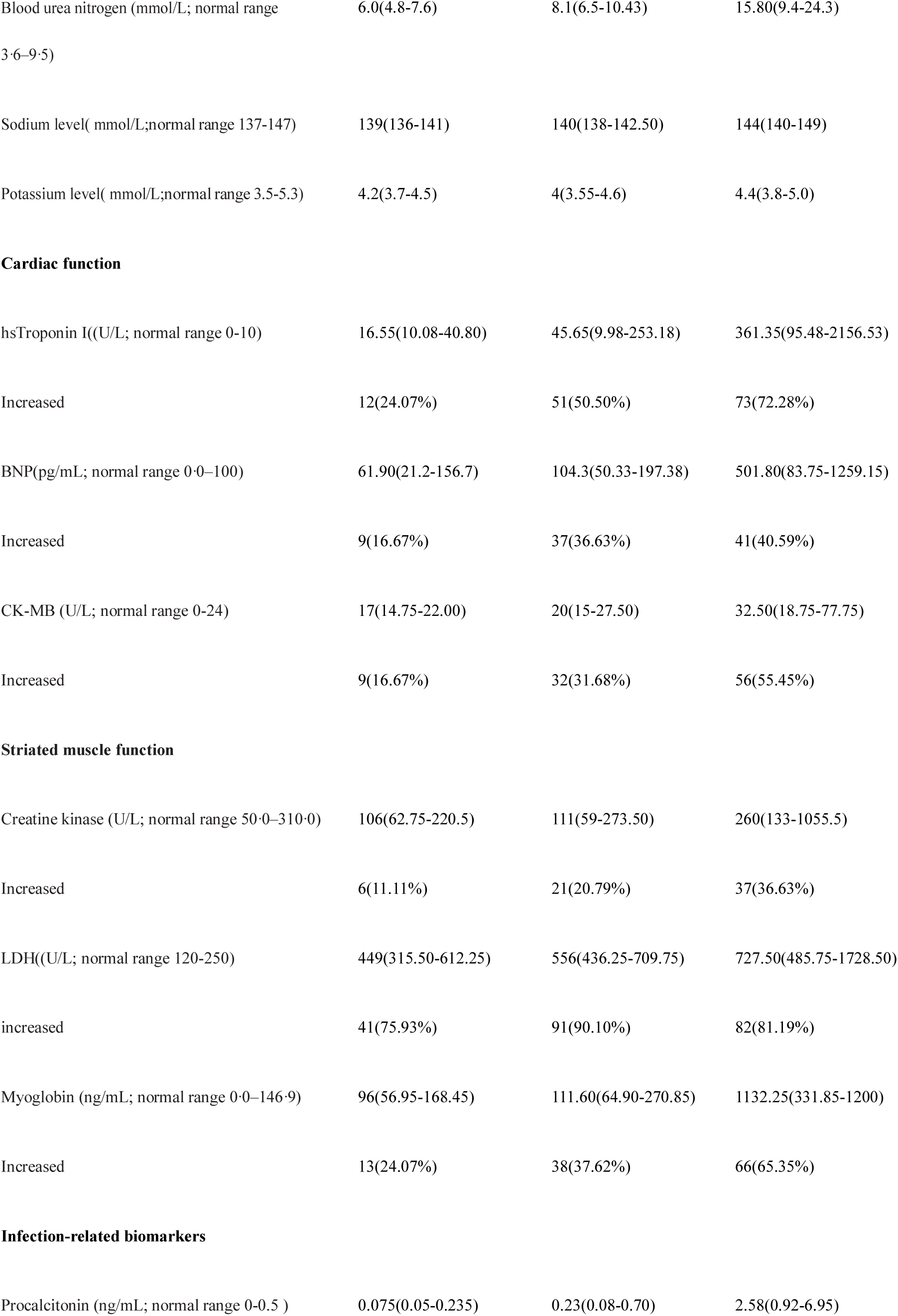
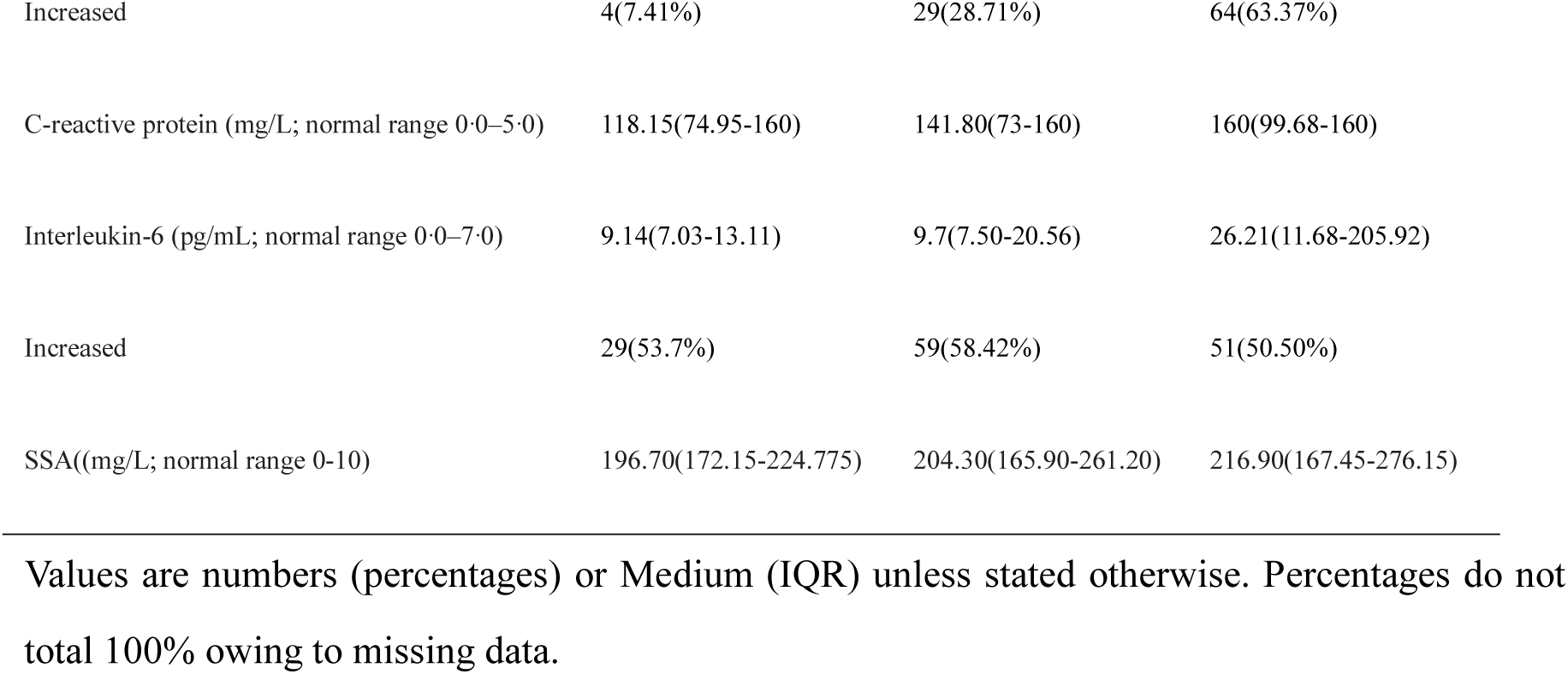
Dynamic Profile of Laboratory Findings in patients with COVID-19

**Table4.** The course of COVID-19,grouped by serum hsTroponin I at admission to general ward

|  | Increased<br>group(N=12) | Normal group(N=28) | P value |
| --- | --- | --- | --- |
| Days from onset to<br>admission | 12(10.25-12.75) | 10(8-12.75) | 0.213 |
| Days from admission<br>to ICU | 1.095(1-3.75) | 6(3-10) | 0.000 |
| Days in ICU | 4(2-8.75) | 5(2-9) | 0.652 |
| Days from onset to<br>death | 18.5(14.5-21.14) | 22.5(19-29.75) | 0.016 |

Values are Medium (IQR) unless stated otherwise. *P* values indicate differences between increased group of hsTroponin I and normal group patients. *P* < .05 was considered statistically significant

**Table5.**
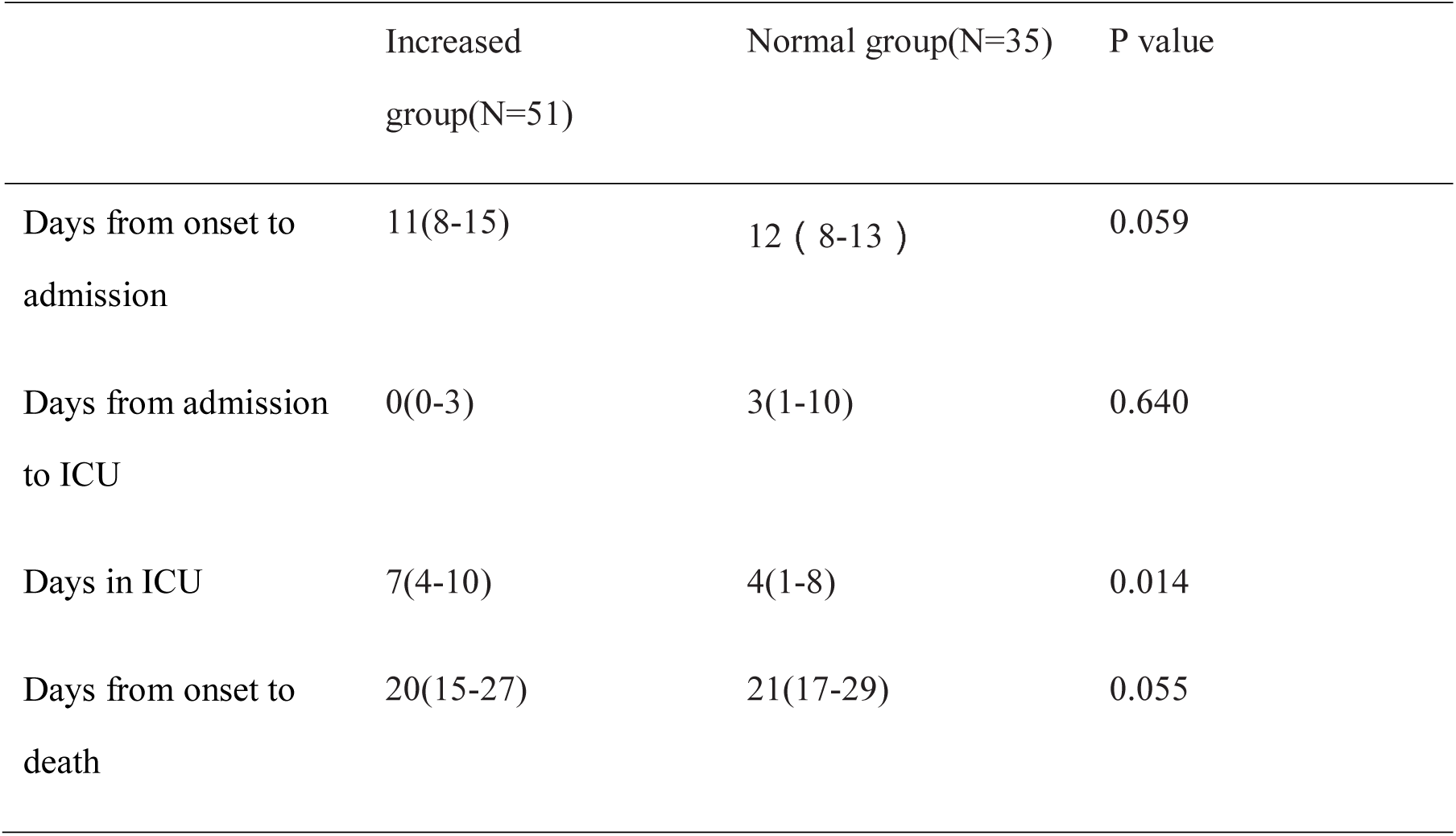
The course of COVID-19,grouped by serum hsTroponin I at admission to ICU

|  | Increased<br>group(N=51) | Normal group(N=35) | P value |
| --- | --- | --- | --- |
| Days from onset to admission | 11(8-15) | 12 ( 8-13 ) | 0.059 |
| Days from admission to ICU | 0(0-3) | 3(1-10) | 0.640 |
| Days in ICU | 7(4-10) | 4(1-8) | 0.014 |
| Days from onset to death | 20(15-27) | 21(17-29) | 0.055 |

### 3.5. Treatments

Ninety seven patients were transferred to the hospital based on laboratory diagnosis or deterioration after treatments in other hospitals. All patients were treated in isolated wards. Among them, 61(60.40%) received antiviral drugs, including Oseltamivir, Ribavirin, Lopinavir, Ritonavir, Ganciclovir, or Interferon, etc. Glucocorticoids, intravenous immunoglobulins, and thymosin preparations were used in, 59(58.42%), 64(63.37%), and 45(44.55%) patients, respectively.. All patients received antibiotic treatment, including cephalosporins and quinolones. More than half of patients received restricted antibiotics(63,62.38%), including carbapenems, linezolid, tigecycline, etc., due to repeated fever or fluctuation of inflammation indicators. 23(22.78%) patients received antifungal drugs.

All patients received oxygen therapy. 84(83.17%) patients were treated with non-invasive ventilator or high-flow oxygen therapy, and 77(76.24%), invasive mechanical ventilation. The median time from ARDS to invasive mechanical ventilation was 3.00 days(IQR0.00-6.00), of which 21 patients were intubated 2 days before deaths. The duration of invasive mechanical ventilation was 1–31 days (median 5 days [IQR2.00-8.00]). Seven patients were treated with Extracorporeal Membrane Oxygenation(ECMO) and eight with Continuous Renal Replacement Therapy(CRRT).

## 4. DISCUSSION

The present analyses revealed the clinical characteristics of 101 deaths caused by COVID-19 in China. Based on our knowledge, this is the largest sample among the reports on the clinical characteristics of dead patients although some studies described a limited number of deaths.

Up to 5^th^ March, 2020, the cumulative number of infected victims was 95,333, and the number of deaths reached 3,282 in many countries of the world. The mortality was up to 3.4%, which is significantly high compared with that of common pneumonia. Among the 101 deaths, males dominated and the proportion of elderly patients with underlying diseases was relatively high. Importantly, we found that the blood group distribution of the deaths (A-42.57%, B-30.69%, O-17.82%, AB-8.91%) remarkably differred from that of Han population in Wuhan^[12]^. Type O was relatively low but Type A was relatively high. ABO blood group antigen substances are widely distributed in the human respiratory, digestive tract, and reproductive systems^[13]^. Previous researches have shown that ABO blood group are related to the onset and spread of various diseases because the blood group antigens may involve in virus infection as receptor^[14-15]^. In the study of various susceptible genes in SARS-CoV, individuals in the blood group O had a lower infection rate^[16]^. Guillon et al. found that type A antibody can provide protection by inhibiting interaction between the virus and ACE2 receptor^[17]^. However, the higher proportion of patients with type A blood remains unclear although the lack of antibody A protection might be involved. Further research is needed to explore the mechanism by which the patients of type A is more susceptibe to COVID-19 infection.

The clinical manifestations of COVID-19 are non-specific, which is consistent with previous studies^[10]^, the most common symptom is fever. However, not all patients had fever, for instance, no fever was noted in 10(9.90%) patients on the onset of disease. Moreover, 21(20.79%) of the patients had no respiratory symptoms in the begining of the disease, and fever and chest tightness gradually occurred with the disease progressed. Therefore, the delay of fever and respiratory symptoms may affect the early identification of COVID-19.

The mortality of critically ill COVID-19 patients is high, but its mechanism is not clear at present, and it may be related to the virus-induced acute lung injury, inflammatory factor storm, multiple organ damage and secondary nosocomial infections. We collected laboratory examination results of patients at admission, at the time of transfer to the ICU, and at 48h before deaths, and found that 71(70.30%) of patients had an elevated level of IL-6 when they were admitted to the hospital, and it gradually increased with the disease progressed and was up to 26.21pg/mL(IQR11.68-205.92) 48h before deaths, significantly higher than results reported by Chen et al^[9]^, indicating a severe inflammatory reaction.We found that 23(22.78%) of patients had abnormal coagulation function at admission, which is mainly manifested as increased D-dimer and a sudden deterioration. Under this circumstance, attention should be paid to the presence of pulmonary thromboembolism after micro-thrombosis in the lungs or deep vein thrombosis. For hypercoagulable patients without contraindications, reasonable anticoagulant therapy is a possible choice.

It has been reported that 2019 novel coronavirus(2019-nCov) can damage function of organs such as the lungs, kidneys, heart, or liver, etc., but no relevant description was seen of the involvement sequence of other organs except lung. We collected indicators of organ damage when patients were admitted to the hospital and found that most of patients had myocardial damage,among them 32 patients were exclusively complicated with myocardial damage. Both 2019-nCov and 2003-SARS belong to the genus β coronavirus, and angiotensin-converting enzyme 2 (ACE2) has been confirmed as a common pathogenic receptor^[19].^ Thirty one percent of ICU patients were found to have significant abnormalities in Troponin I(TNI) ^[8]^. We noted that TNI was significantly elevated in the ICU patients or before deaths compared with primary admission to hospital. Further analysis revealed that patients who were hospitalized with elevated troponin underwent a fast progress from onset to ICU and to death. The results suggest that heart is a potential targeted organ by 2019-nCOV in the disease process. Thus monitoring and prediction of heart injury would be needed at early stage as well as in the course of disease.

Additionally, we found that with the disease progressed, counts of leukocytes, neutrophils, PCT and CRP level gradually increased, while lymphocytes increased again after the decline, and some patients experienced a drop of body temperature and then increased or continued fever fluctuations. These indicators do not fully meet the characteristics of viral infections. It is necessary to be alert to that patients who may have hospital-acquired pneumonia, especially bacterial infections. The sputum culture results also indicated bacterial and fungal infections, such as *Klebsiella pneumoniae, Acinetobacter baumannii* and *Candida albicans* in some patients. Secondary bacterial and fungal infections may be related to suppressed immunity, lack of medical resources and unsmooth sputum drainage. In addition, clinical manifestations of bacterial or fungal infections in critical patients may be unconspicuous due to their compromised immunity or hormone therapy. Therefore, it is essential to keep monitoring temperature, laboratory indicators, imaging indicators and airway secretion characteristics of the patients. Prevention and control of nosocomial infections also needed.

In terms of treatment, usage of antiviral drugs and hormones and related course is still controversial. In this study, 60.40% of patients received antiviral drugs, 58.42% patients were given glucocorticoids, and the treatment course was mostly 3-5 days. All patients received antibiotic treatments including cephalosporins and quinolones. More than half of patients received restricted antibiotics due to fluctuations of body temperature or inflammation indicators. Mechanical ventilation is the main supportive treatment for critically ill patients, but the overall survival time of patients after invasive mechanical ventilation was short(median 5 days [IQR2.00-8.00]), and most patients did not benefit significantly from invasive mechanical ventilation and had suffered multiple organ failure caused by severe hypoxia before invasive mechanical ventilation. Therefore, for critical patients, early invasive mechanical ventilation treatment should be in consideration.

This study has some limitations. Only the critical death patients are included, and no comparison was made between the improvement groups. The sample size can be increased in further research for prospective case-control study. The blood group composition of patients cannot be statistically analyzed and whether the blood group difference is related to the susceptibility of COVID-19 infection needs to be clarified.

## 5. CONCLUSION

Critical COVID-19 may cause fatal respiratory distress syndrome and multiple organ failure with high mortality rate.The blood group distribution of the deaths remarkably differred from that of Han population in Wuhan.Heart may be the earliest damaged organ except the lungs. Hospital-acquired pneumonia in the later period is worthy of attention.

## Data Availability

This study collected Death Cases with COVID-19 from Dec 30, 2019 to Feb 16, 2020 in Intensive care unit(ICU) of Wuhan Jinyintan Hospital.This study is approved by the Ethics Committee of Wuhan Jinyintan Hospital Approval, and all relevant personnel exempt from informed consent due to the particularity of the disease outbreak.

